# The physio-affective symptoms of Long COVID are strongly predicted by the severity of the acute infectious phase, and lowered antioxidants, nitric oxide, and alanine transaminase levels

**DOI:** 10.1101/2025.02.07.25321870

**Authors:** Michael Maes, Chavit Tunvirachaisakul, Laura de Oliveira Semeão, Ana Paula Michelin, Andressa K. Matsumoto, Francis F. Brinholi, Decio S. Barbosa, Yingqian Zhang, Pimpayao Sodsai, Nattiya Hirankarn, Abbas F. Almulla

## Abstract

**Aims:** This study investigated a) whether the affective, chronic fatigue syndrome/myalgic encephalomyelitis (CFS/ME), respiratory, and neurological Long COVID (LC) symptom domains are intercorrelated manifestations of an underlying construct, namely the LC phenome; and b) the predictive effects of clinical symptoms and biomarkers of the COVID-19 infectious phase on LC.

**Methods:** This cohort study included 71 LC patients who had suffered from COVID-19 six months prior. The patients complied with the LC disease criteria set forth by the World Health Organization (WHO). We used the WHO symptom list to score the LC symptoms and assessed SARS-CoV-2 positivity using real-time reverse transcription-polymerase chain reaction (RT-PCR), white blood cell counts, anion gap, alanine transaminase (ALT) levels, paraoxonase 1 (PON1) activity and genotypes, total radical trapping potential of plasma (TRAP), and nitric oxide metabolites (NOx).

**Results:** All four clinical LC subdomains are significantly intercorrelated, allowing for the extraction of a single latent construct encompassing affective, CFS/ME, respiratory, and neurological symptoms, labeled the physio-affective phenome of LC. The latter was strongly predicted by dyspnea and productive sputum, RT-PCR SARS-CoV-2 cycle threshold values, lower anion gap, and increased neutrophil percentage during the infectious phase. Other predictive biomarkers were lower ALT, TRAP, and NOx levels, and the PON1 QR192 over dominant gene model (protective).

**Conclusions:** LC may be a clinically homogeneous disease characterized by highly intercorrelated affective, CFS, neurological, and respiratory subdomains. The physio-affective phenome and its domains are strongly predicted by biomarkers of acute infection and associated inflammation and oxidative stress.

## Introduction

Following the COVID-19 pandemic, which led to millions of acute cases, healthcare professionals now face a further challenge arising from the emergence and/or persistence of symptoms that endure beyond the acute infectious phase of severe acute respiratory syndrome coronavirus (SARS-CoV-2) infection (Fernández-de-Las-Peñas et al., 2021). The condition is commonly known as Long COVID (LC), Long coronavirus disease 2019, post-coronavirus disease 2019, or post-COVID-19. This condition is a sequel to prior infection with SARS-CoV-2 and is generally marked by the continuation of symptoms that extend beyond a three-month period following the acute phase of COVID-19 (Greenhalgh et al., 2024; Nalbandian et al., 2021; Tan et al., 2024; World Health Organization, 2022).

This syndrome presents itself as a constellation of symptoms, predominantly including, though not exclusively limited to affective symptoms (including depressed mood, loss of interest, anxiety, loss of appetite, weight loss, impaired decision making), chronic fatigue syndrome (CFS) (including chronic fatigue and fibromyalgia) and myalgic encephalomyelitis (including post-exertional malaise, PEM), respiratory symptoms (dry cough, dyspnea, sputum), neurological symptoms (including tremor, loss of taste and smell, dizziness, numbness, unsteady gait), and gastro-intestinal symptoms (including constipation, diarrhea, stomach pain) (Renaud-Charest et al., 2021; Sandler et al., 2021; Titze-de-Almeida et al., 2022; World Health Organization, 2022). Nonetheless, the symptoms that are most frequently documented in association with long-term COVID encompass CFS/ME, affective symptoms such as anxiety and depression, cognitive deficits, and respiratory issues (Bliddal et al., 2021; Goldenberg, 2024; Mazza et al., 2022; Phu et al., 2023; van den Borst et al., 2021). For instance, within a period of six months following the onset of the first COVID-19 symptom, nearly one-third of individuals who have recovered from the infection indicate that they encounter neuropsychiatric symptoms, such as insomnia, anxiety, or depressive episodes (Taquet et al., 2021).

The National Institute for Health and Care Research (NIHR) has posited that LC may encompass various syndromes, including post-intensive care syndrome (PICS), long-term COVID syndrome, and post-viral fatigue syndrome (Mahase, 2020). Some scholars argue that LC can present in various forms, encompassing one or more identifiable medical conditions (Ely et al., 2024; NIHR, 2025). Nonetheless, the laboratory of the primary author of the present study has indicated that affective symptoms, anxiety, symptoms associated with CFS/ME, and physio-somatic symptoms exhibit a significant interconnection during the course of LC, to the degree that a single latent construct can be derived (Al-Hakeim et al., 2023a; Al-Hakeim et al., 2023b; Almulla et al., 2024d; Maes et al., 2024). Consequently, this latent factor that underpins the symptoms of LC has been designated as the physio-affective phenome of LC.

The intensity of the physio-affective phenome associated with LC is significantly predicted by the severity of the acute phase of SARS-CoV-2 infection, as evidenced by elevated peak body temperature (PBT) and diminished oxygen saturation (SpO2) levels (Al-Hadrawi et al., 2022). Nonetheless, it remains unclear whether the symptoms observed during the acute infectious phase, the cycle threshold (Ct) values for SARS-CoV-2 as determined by real-time PCR, and the biomarkers associated with the acute phase of SARS-CoV-2 infection can serve as predictors for the severity of LC or its symptomatic subdomains. During the acute infectious phase of SARS-CoV-2 infection, one observes elevated white blood cell (WBC) counts, along with an increase in the number or percentage of neutrophils (Castanheira and Kubes, 2023). Moreover, acid-base imbalances have been documented in individuals suffering from acute SARS-CoV-2 infections, encompassing conditions such as metabolic and respiratory acidosis, as well as metabolic alkalosis (Al-Azzam et al., 2023; Zemlin et al., 2022).

It is also well established that infection with SARS-CoV-2 may lead to liver damage, as evidenced by elevated levels of alanine transaminase (ALT) (Sanyaolu et al., 2023). Recent findings indicate a correlation between heightened oxidative stress, characterized by lipid peroxidation and nitric oxide production, diminished antioxidant levels, and the concomitant activation of immune-inflammatory responses with the physio-affective phenome observed in LC (Al-Hakeim et al., 2023a; Al-Hakeim et al., 2023b). Affective symptoms frequently coincide with a reduction in the total radical trapping antioxidant potential of plasma (TRAP) and the activities of paraoxonase 1 (PON1) (Maes et al., 2021). Nonetheless, it has yet to be determined whether modifications in these biomarkers associated with the acute infectious phase predict the severity of LC.

Hence, the current investigation was undertaken to explore the extent to which the affective, CFS/ME, respiratory, gastrointestinal, and neurological symptomatic domains of LC exhibit strong intercorrelations and whether they represent manifestations of a shared latent trait. This study further seeks to investigate the potential predictive relationship between the symptoms of LC and various biomarkers observed during the acute infectious phase. These include white blood cell counts, the percentage of neutrophils, acid-base imbalances, ALT levels, as well as diminished PON1 activity and TRAP levels, and elevated nitric oxide metabolite (NOx) production.

## Patients and Methods

### Patients

This is a retrospective cohort study that enrolled 71 patients with LC who had previously experienced acute COVID-19 infection. All patients were enrolled at the Department of Microbiology and Immunology, Chulalongkorn University, Bangkok, Thailand. We initially enrolled 98 LC patients in the study; however, six participants did not meet all the criteria of LC symptoms, seven others did not participate in longitudinal clinical data collection, and blood samples from 14 participants were unavailable for analysis. Expert virologists and physicians diagnosed both the acute phase of COVID-19 infection and LC. All subjects were diagnosed during the acute infectious phase using the typical acute infectious symptoms, a positive SARS-CoV-2 reverse transcription real-time polymerase chain reaction (rRT-PCR) test, and the identification of immunoglobulin (Ig)G and IgM antibodies against SARS-CoV-2. The patients were selected according to the LC disease criteria set forth by the World Health Organization (World Health Organization, 2022). The criteria encompassed: a) individuals with a confirmed acute COVID-19 infection; b) individuals exhibiting a minimum of two symptoms identified by the WHO criteria; c) symptoms persisting for no less than 3 months; d) symptoms extending beyond the acute phase of SARS-CoV-2 infection or manifesting 2-3 months post-initial infection; and e) symptoms remaining evident 6 months following the acute phase. The symptoms are enumerated in the Electronic Supplementary File (ESF), Table 1. The symptom domains encompass affective symptoms, CFS/ME symptoms, neuronal symptoms, respiratory symptoms, and gastro-intestinal symptoms. The overall severity of LC was defined as the first principal component derived from the domains of affective symptoms, CFS/ME symptoms, neurological symptoms, and respiratory symptoms.

**Table 1.** Demographic and clinical data of the Long COVID patients divided into two groups using cluster analysis.

| Variables | Cluster 1<br>HC (n=43) | Cluster 2<br>(n=28) | F/$\chi^2$ | df | p |
| --- | --- | --- | --- | --- | --- |
| Age (years) | 40.9 (13.1) | 42.6 (11.2) | F=0.33 | 1/69 | 0.565 |
| Sex (Female/Male) | 19/24 | 21/7 | $\chi^2=6.55$ | 1 | 0.011 |
| Education (ISCO score) | 4.72 (2.17) | 4.73 (2.00) | F=0.00 | 1/69 | 0.988 |
| BMI (kg/m<sup>2</sup>) | 26.30 (4.78) | 25.15 (4.00) | F=1.11 | 1/69 | 0.296 |
| Acute SARS-CoV-2 infectious symptoms (No/Yes) | 36/7 | 13/15 | $\chi^2=11.03$ | 1 | 0.001 |
| Depression score* | 0.80 (0.26) | 4.36 (0.33) | F=69.29 | 1/66 | <0.001 |
| CFS/ME score* | 0.43 (0.11) | 1.92 (0.14) | F=71.15 | 1/66 | <0.001 |
| Respiratory symptom score* | 0.09 (0.10) | 1.27 (0.12) | F=56.69 | 1/66 | <0.001 |
| Neuronal symptom score* | 0.26 (0.17) | 2.02 (2.21) | F=41.32 | 1/66 | <0.001 |
| GIS symptom score* | 0.04 (0.05) | 0.24 (0.06) | F=6.56 | 1/66 | 0.013 |
| Overall severity of Long COVID* | -0.640 (0.098) | 1.019 (0.125) | F=104.88 | 1/66 | <0.001 |

Results are shown as mean ( ± SD). FEPT: Fisher's exact probability test; $\chi^2$ : analysis of contingency tables, F: results of analysis of variance, \* Results of univariate GLM analysis with age and sex as covariates; shown as marginal estimated means (SE).
BMI: body mass index; CFS/ME: chronic fatigue syndrome / myalgic encephalomyelitis, GIS: gastro-intestinal.

This cohort study was conducted to investigate the impact of symptoms and biomarkers of acute SARS-CoV-2 infection on LC symptoms. In the acute phase of SARS-CoV-2 infection, we evaluated the existence of dyspnea and productive sputum (No/Yes) as indicators of the acute SARS-CoV-2 infectious phase. The indicators of the acute infectious phase were total white blood cell (WBC) counts, the percentage of neutrophils and lymphocytes, their ratio (N/L ratio), serum ALT or glutamate-pyruvate transaminase (SGPT), and the anion gap. Three weeks post-acute phase, we evaluated serum indicators of oxidative and nitrosative stress.

We excluded all patients who had previously suffered significant depression, dysthymia, CFS/ME, schizophrenia, bipolar disorder, psycho-organic syndromes, substance use disorders (except nicotine dependency), panic disorder, and generalized anxiety disorder prior to the acute infection phase. Furthermore, we excluded all individuals with systemic immunological or autoimmune illnesses, including scleroderma, psoriasis, systemic lupus erythematosus, chronic obstructive pulmonary disease, chronic kidney disease, inflammatory bowel disease, and rheumatoid arthritis. We additionally excluded pregnant or lactating women.

The Institutional Review Board of Faculty of Medicine, Chulalongkorn University in Bangkok, Thailand has authorized the conduct of this research (IRB number 114/64). Before inclusion in the study, all individuals or their legal representatives provided written informed consent. The study’s design and execution adhered to the criteria established by the International Conference on Harmonization of Good Clinical Practice, the Belmont Report, the Council of International Organizations of Medical Sciences (CIOMS) Guidelines, in addition to ethical and privacy regulations in Iraq and internationally. Our institution’s review board complies with the International Guidelines for the Conduct of Safe Human Research (ICH-GCP).

### Methods

Approximately 6 months following the acute infectious period, we evaluated all symptoms of LC as either present or absent, as illustrated in Electronic Supplementary File (ESF), Table 1. The sum of assorted items across the five subdomains was calculated, resulting in scores for affective symptoms (score: 0-10), CFS/ME symptoms (score: 0-5), neurological symptoms (score: 0-14), respiratory symptoms (score: 0-4), and gastrointestinal symptoms (score: 0-4).

Fasting blood samples for the investigation of biomarkers associated with active SARS-CoV-2 infection were collected during the acute infectious phase. The anion gap was computed using the formula: the total sodium and potassium concentrations minus the total chloride and bicarbonate concentrations. The levels of sodium, potassium, and chloride were assessed utilizing an ion-selective electrode dilution (indirect) technique. Carbon dioxide, indicative of bicarbonate concentrations, was quantified enzymatically by PEP carboxylase. The studies were performed using the Alinity C analyzer (Abbott Laboratories, USA), produced in Tochigi-ken, Japan. The coefficients of variation (CV) for these tests were 0.9%, 1.3%, 1.1%, and 5.5%, respectively. ALT levels were assessed with a NADH-based technique devoid of P-5’-phosphate. The experiment was conducted using the identical Alinity C analyzer, exhibiting a coefficient of variation of 4.1%. The WBC count, together with neutrophil and lymphocyte percentages, was assessed using the XN-10 analyzer produced by Sysmex in Kobe, Japan. The analysis was performed by flow cytometry with a semiconductor laser. The coefficients of variation (CV) for total WBC count, neutrophils, and lymphocytes were 2.1%, 2.3%, and 3.5%, respectively. The measurement of cycle threshold (Ct) values for SARS-CoV-2 was conducted utilizing the Cobas® SARS-CoV-2 assay on the Cobas® 6800/8800 system (Roche Diagnostics, Basel, Switzerland), produced in Rotkreuz, Switzerland. This real-time RT-PCR test is intended to identify two target areas of the SARS-CoV-2 genome: the ORF1ab gene, specific to SARS-CoV-2, and the E gene, serving as a pan-sarbecovirus marker. The method utilizes entirely automated procedures, encompassing sample preparation, amplification, and detection, thereby guaranteeing elevated sensitivity and specificity. Nasopharyngeal or oropharyngeal swabs obtained in viral transport media were handled in accordance with the manufacturer’s protocol. The system software automatically generated Ct values for each target gene, reporting the coefficient of variation (% CV) for the ORF1ab and E gene targets as 0.89% and 1.27%, respectively, which signifies great precision in the assay’s performance. We additionally categorized the SARS-CoV-2 Ct values using a threshold of ≤ 23 Ct (designated as 1, whereas Ct > 23 is designated as 0).

Fasting blood for the assessment of oxidative stress indicators was collected at 8:00 a.m. after an overnight fast, three weeks post-acute infection phase. Serum and plasma aliquots were stored at −80 °C until thawed for analysis. We evaluated total TRAP, NOx, the PON1 genotypes, and the PON1 CMPAase and AREase activities. TRAP was quantified by chemiluminescence, as outlined by (Repetto et al., 1996). The 2,2’ azo-bis swiftly produces peroxyl radicals by its interaction with carbon-centered radicals and molecular oxygen. These free radicals interact with luminol, which functions as an amplifier, resulting in chemiluminescence. The incorporation of plasma diminishes baseline chemiluminescence for a duration (induction time) that correlates with the concentration of TRAP, until free radicals are replenished, restoring starting chemiluminescence levels. The experiment was performed using a Beckman β counter, model LS6000 (Fullerton, CA, USA), in a non-coincident counting mode for 25 minutes, with a response range of 300 to 620 nm. NOx were quantified by assessing nitrite and nitrate concentrations using a microplate reader (EnSpire®, Perkin Elmer, USA) at 540 nm (Navarro-Gonzálvez et al., 1998), with results expressed in molarity (M). All inter-assay coefficients of variation were less than 10.0%. The PON1 status was assessed using AREase and CMPAase activity, as well as PON1 Q192R genotypes in the current study (Brinholi et al., 2023; Maes et al., 2022; Matsumoto et al., 2021; Moreira et al., 2019a; Moreira et al., 2019b). We examined the hydrolysis rate of phenylacetate at low salt concentrations by assessing the activity of AREase and CMPAase (Sigma, St. Louis, MO, USA) (Brinholi et al., 2023; Maes et al., 2022; Matsumoto et al., 2021; Moreira et al., 2019a; Moreira et al., 2019b). A Perkin Elmer® EnSpire model microplate reader (Waltham, MA, USA) was employed to assess the rate of phenylacetate hydrolysis at a constant temperature of 25 °C during a duration of 4 minutes, comprising 16 measurements taken at 15-second intervals. The activity was quantified in units per milliliter (U/mL) using the phenyl acetate molar extinction coefficient of 1.31 mMol/L cm-1. CMPA and phenyl acetate were employed to categorize the functional genotypes of the PON1Q192R polymorphism (PON1 192Q/Q, PON1 192Q/R, and PON1 192R/R) (Sigma, PA, USA). The phenylacetate reaction is performed at elevated salt concentrations, which partially impedes the activity of the R allozyme, facilitating a clearer differentiation among the three functional genotypes. We examined the dominant, recessive, additive, and over dominant models of the PON1 genotype.

#### Statistics

The statistical analysis for this study was conducted utilizing version 28 of IBM SPSS for Windows. Pearson’s product-moment correlation coefficients were utilized to ascertain statistical correlations among continuous variables. The researchers employed analysis of variance to investigate the relationships between diagnostic categories and clinical data. The χ^2^-test or Fisher’s exact probability test were utilized by the researchers as a contingency table analysis to evaluate the statistical associations between categorical variables. Univariate GLM analysis was utilized to elucidate the relationships between clinical and biomarker data and the LC groups identified through cluster analysis. These findings were controlled, if needed, for confounding variables, including age, sex, BMI, and education years. Manual and stepwise automatic multiple linear regression analyses were utilized to investigate the impact of biomarkers on the severity of LC subdomains. The automatic forward stepwise regression method employed a p-to-enter threshold of p=0.05, while a p-value of 0.06 was utilized for the exclusion of variables from the final regression analysis. The model statistics included F statistics and corresponding p-values, while the effect size was evaluated using R^2^, specifically the partial Eta squared. Additionally, standardized β coefficients and t-statistics (along with p-values) were calculated for each variable used in the final regression models. The White test and a modified version of the Breusch-Pagan test were conducted to determine the presence of heteroskedasticity. We analyzed the data for potential multicollinearity and collinearity, employing a tolerance limit of 0.25 and a variance inflation factor threshold of four or higher. Furthermore, for this research, we utilized the condition index and variance proportions derived from the collinearity diagnostics table. A binary logistic regression analysis (oversampling method) was conducted to identify the biomarkers that most accurately predicted severe versus non-severe LC syndrome. We calculated the odds ratio with 95% confidence intervals, as well as the accuracy and Nagelkerke metrics, which served as the effect size. We employed a stepwise automatic and manual approach to identify the optimal predictors. We employed a p-value for entry of 0.05 and a p-value for removal of 0.06. As required, we employed transformations like logarithmic, square-root, and rank-based inverse normal to guarantee that our data indicators conformed to a normal distribution. All analyses were conducted using two-tailed tests, with a p-value (alpha level) of 0.05 or lower being statistically significant. Principal component analysis was employed as a feature reduction technique to investigate the extraction of the first principal component from various LC symptom domains. The first principal component derived from the data must adhere to stringent criteria: a) all loadings on this component must exceed 0.7; b) the first principal component must account for over 50% of the variance; and c) the Kaiser-Meyer-Olkin measure must surpass 0.7, with Bartlett’s test of sphericity yielding significant results. Two-Step cluster analysis was employed to identify potential clusters within the dataset of symptomatic LC domains. We employed log-likelihood as the distance metric and the Schwarz-Bayesian Criterion as the grouping criterion. The criterion for accepting the cluster solution was that the average silhouette of cohesiveness and separation exceeded 0.5.

The principal statistical analysis employed was the multiple regression analysis with the overall severity of LC as the dependent variable and the biomarkers and acute SARS-CoV-2 pneumonia symptoms as explanatory variables. G*Power 3.1.9.4 showed that the a priori anticipated sample size was n=58, utilizing an explained variance of approximately 15% and thus an effect size of f^2^ = 0.25, with an alpha of 0.05, power of 0.8, and a maximum of 5 predictors.

## Results

### Cluster analysis

To examine whether relevant clusters in the LC data set could be retrieved we performed a two-step cluster analysis on depression, CFS/ME, respiratory, neurological, and GIS symptoms. We found that a two-step cluster solution generated two adequate clusters with a cluster size ratio of 1.54 and an average silhouette measure of cohesion and separation of 0.6. The most important predictor was CFS/ME (predictor importance of 1.0) followed by depression symptoms (0.92), respiratory symptoms (0.87), neurological symptoms (0.71) and GIS symptoms (0.32). **Table 1** shows that the first cluster comprised 43 subjects and the second cluster 28 subjects. There were no significant differences in age, education level, and BMI between both clusters. There were significantly more women in cluster 2 than in cluster 1. The 5 LC symptom domains were significantly higher in cluster 2 than in cluster 1. The effect sizes were in descending order of importance: CFS/ME (0.506), depression (0.492), lung (0.453), neurological (0.370) and GIS (0.081). The frequency of the acute SARS-CoV-2 respiratory symptoms was significantly higher in cluster 2 than in cluster 1.

### Results of PCA

The depression score was significantly correlated wth the CFS/ME (r=0.742, p<0.001), neurological (r=0.673, p<0.001) and respiratory (r=0.625, p<0.001) scores. The CFS/ME score was significantly associated with neurological (r=0.672, p<0.001) and respiratory (r=0.640, p<0.001) scores. There was also a significant correlation between the neurological and respiratory scores (r=0.674, p<0.001). The GIS symptom score was significantly associated with the depression (r=0.511, p<0.001), CFS/ME (r=0.394, p<0.001), neurological (r=0.418, p<0.001) and respiratory (r=0.373, p<0.001) scores.

**Table 2** shows the results of principal component analysis. The first analysis (PCA1 in Table 2) shows that one principal componenet could be extracted from the depression, CFS/ME, respiratory, and neurological symptom scores. The KMO value was greater than 0.65, all loadings were higher than 0.843 and the first principal component explained 73.346% of the variance. The second analysis (PCA2 in Table 2) shows that the GIS symptom score loading was 0.626. Nevertheless, we computed the principal component score extracted from depression, CFS/ME, respiratory, and neurological symptom scores since GIS symptoms were somewhat less strongly associated with the first principal component. Table 1 shows that this index was significantly higher in cluster 2 than in cluster 1.

**Table 2.** Results of principal component analysis (PCA).

| **Variables** | **PCA1**<br>**Loadings** | **PCA2**<br>**Loadings** |
| --- | --- | --- |
| Depression score | **0.877** | **0.882** |
| CFS/ME score | **0.881** | **0.862** |
| Respiratory symptom score | **0.843** | **0.823** |
| Neurological symptom score | **0.870** | **0.856** |
| GIS symptom score | - | **0.626** |
| KMO | 0.829 | 0.849 |
| Bartlett $\chi^2$ | 155.411 | 176.155 |
| P value | df=6, p<0.001 | df=10, p<0.001 |
| Explained variance | 73.346% | 66.490% |

CFS/ME: chronic fatigue syndrome / myalgic encephalomyelitis; GIS: gastro-intestinal; KMO: Kaiser-Meyer-Olkin metric

### Biomarker measurements in Long COVID

**Table 3** shows the measurements of the biomarkers in both cluster analysis-generated groups. After FDR p correction, the NOx levels remained significantly different between the two groups with lower levels in cluster 2. The other biomarkers did not significantly differ between both groups. **Table 4** lists the results of binary logistic regression analysis with cluster 2 as dependent variable. Table 4, regression #1 shows that cluster 2 was best predicted by lowered NOx and TRAP values and increased severity of acute SARS-CoV-2 pneumonia symptoms and lowered PCR SARS-CoV-2 Ct values, dichotomized using a cut off value of ≤ accuracy was 72.4% (χ^2^=31.66, df=4, p<0.001, Nagelkerke=0.371). We have rerun the same analysis but now without acute SARS-CoV-2 symptoms. This binary logistic regression (regression #2) shows that cluster 2 was best predicted by the combined effects of lowered Nox and PON1 genotype 2 (both inversely associated) and by female sex (χ^2^=26.98, df=3, p<0.001, Nagelkerke=0.320, accuracy=72.7%). PEM was best predicted (regression #3) by the combined effects of TRAP (inversely associated) and by acute SARS-CoV-2 pneumonia symptoms and the dichotomized PCR SARS-CoV-2 values (both positively associated) (χ^2^=25.91, df=3, p<0.001, Nagelkerke=0.311, accuracy=69.4%).

**Table 3.** Baseline biomarkers and oxidative stress biomarkers three weeks after the acute infectious COVID-19 phase in patients with severe (cluster 2) versus mild (cluster 1) Long COVID.

| Variables | Cluster 1<br>HC (n=43) | Cluster 2<br>(n=28) | F/$\chi^2$ | df | p |
| --- | --- | --- | --- | --- | --- |
| White blood cell count ($10^9/L$) | 5277 (255) | 5748 (310) | F=1.29 | 1 | 0.260 |
| Neutrophil % | 55.6 (2.0) | 61.0 (2.5) | F=2.69 | 1/61 | 0.106 |
| Lymphocytes % | 33.6 (1.9) | 29.5 (2.3) | F=1.82 | 1/61 | 0.182 |
| Neutrophil / Lymphocyte ratio | 2.10 (0.30) | 2.92 (0.37) | F=2.74 | 1/61 | 0.103 |
| PCR SARS-CoV-2 (cycle threshold) | 26.35 (0.72) | 24.80 (0.87) | F=1.76 | 1/61 | 0.189 |
| ALT (U/L) | 28.56 (2.50) | 28.64 (2.43) | F=0.00 | 1/61 | 0.980 |
| Anion gap (meq/L) | 14.08 (0.31) | 14.08 (0.31) | F=0.03 | 1/61 | 0.868 |
| PON1 QR192 genotype (QQ/QR/RR) | 5/26/12 | 6/9/13 | $\chi^2=5.46$ | 2 | 0.065 |
| CMPAase (U/mL) | 32.37 (1.26) | 34.03 (1.53) | F=0.66 | 1/61 | 0.419 |
| AREase (U/mL) | 278.45 (6.34) | 278.70 (7.71) | F=0.00 | 1/61 | 0.981 |
| Total radical trapping potential ($\mu M$ Trolox/mg/dL) | 932.39 (63.47) | 986.58 (77.20) | F=0.42 | 1/61 | 0.521 |
| Nitric oxide metabolites ($\mu mol/L$) | 5.11 (0.27) | 3.71 (0.33) | F=10.30 | 1/61 | 0.002 |

Results are shown as mean ( ± SD). $\chi^2$ : analysis of contingency tables, F: results of analysis of variance.
PCR SARS-CoV-2: cycle threshold values for SARS-CoV-2 utilizing real-time reverse transcription-polymerase chain reaction; ALT: alanine aminotransaminase; PON1: paraoxonase 1; CMPAase and AREase: two different PON1 activities.

**Table 4.**
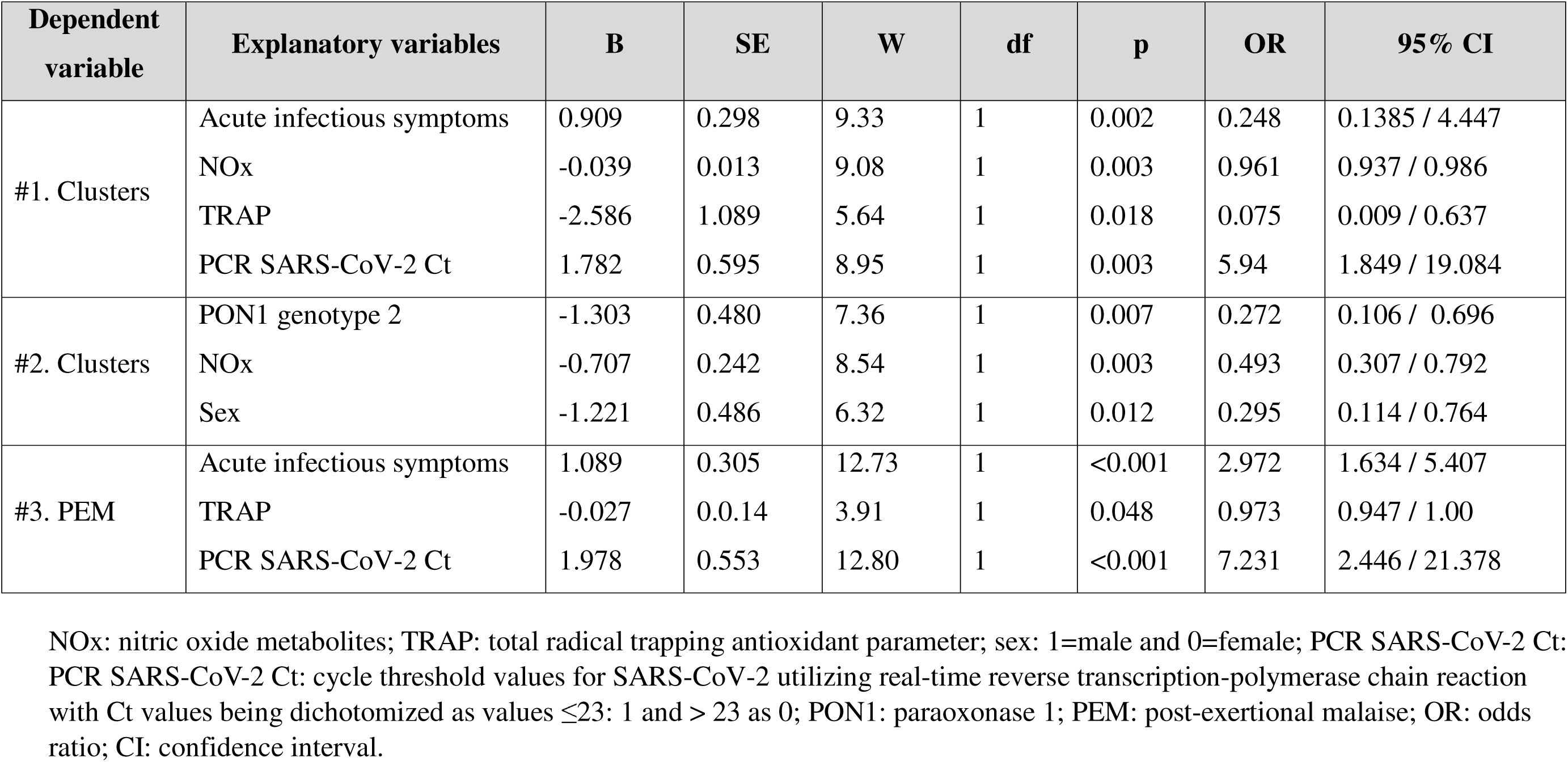
Results of logistic regression analyses with symptom domains as dependent variables.

### Results of multiple regression analysis

**Table 5** shows the results of multiple regression analyses with the LC symptom domain scores as dependent variables and the biomarkers as explanatory variables, whilst allowing for the effects of age, sex, and education. Regression #1 shows that 39.6% of the variance in the overall severity of LC was explained by acute SARS-CoV-2 symptoms (positively), NOx and the quantitative PCR Ct values (both inversely). **Figure 1** shows the partial regression of the overall severity of LC on the severity of COVID-19 infection symptoms. **Figure 2** shows the partial regression of the overall severity of LC on the PCR Ct values. We found that 42.1% of the variance in depression symptoms (regression #2) was explained by acute infectious symptoms (positive association), ALT, anion gap, and the quantitative PCR Ct values (all three inversely associated). After deleting the acute infectious symptoms and the quantitative Ct scores (regression #3), we found that 24.5% of the variance in the depression scores was explained by NOx, anion gap (both inversely) and increased neutrophil%. A large part of the variances in the CFS/ME score (regression #4, 38.6%) and respiratory symptom (regression #5, 28.7%) was explained by acute infectious symptoms and the quantitative SARS-CoV-2 Ct values. In addition, regression #6 shows that the neurological symptoms were best predicted by the acute infectious symptoms (positively), the quantitative SARS-CoV-2 Ct values and ALT (both positively). A larger part of the variance (20.2%) in acute infectious symptoms was explained by the WBC count.

**Table 5.**
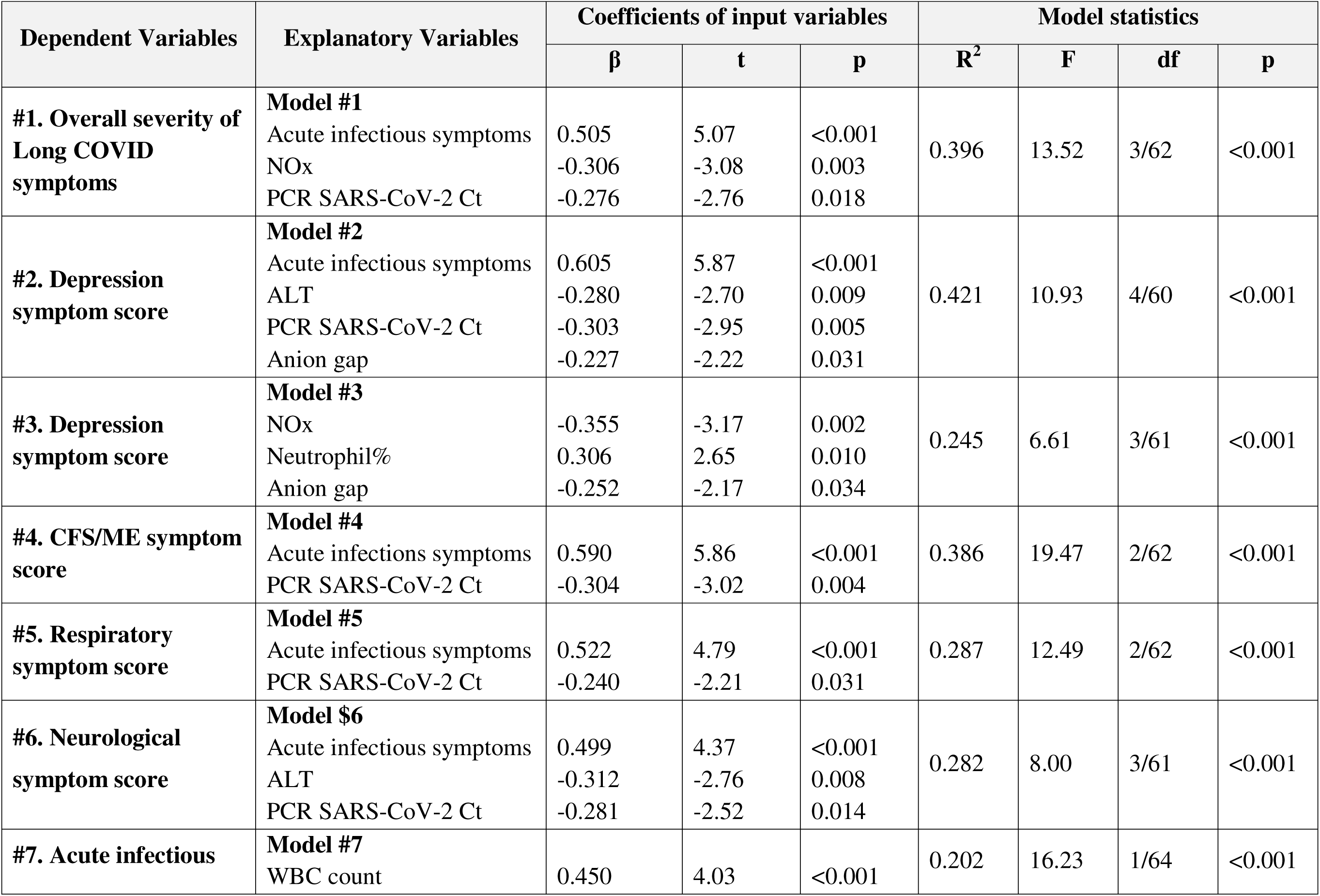
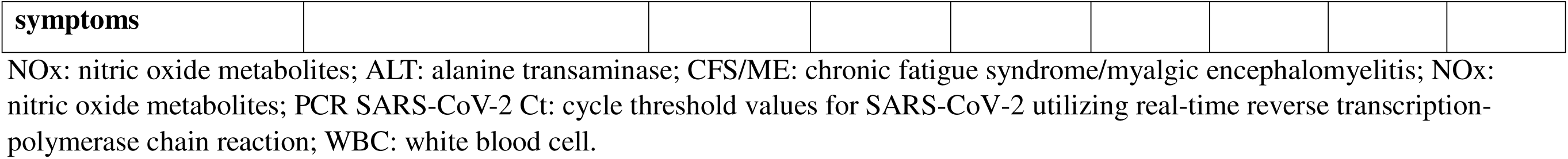
Results of multiple regression analysis with clinical rating scores of Long COVID disease as dependent variables.

**Figure 1.**
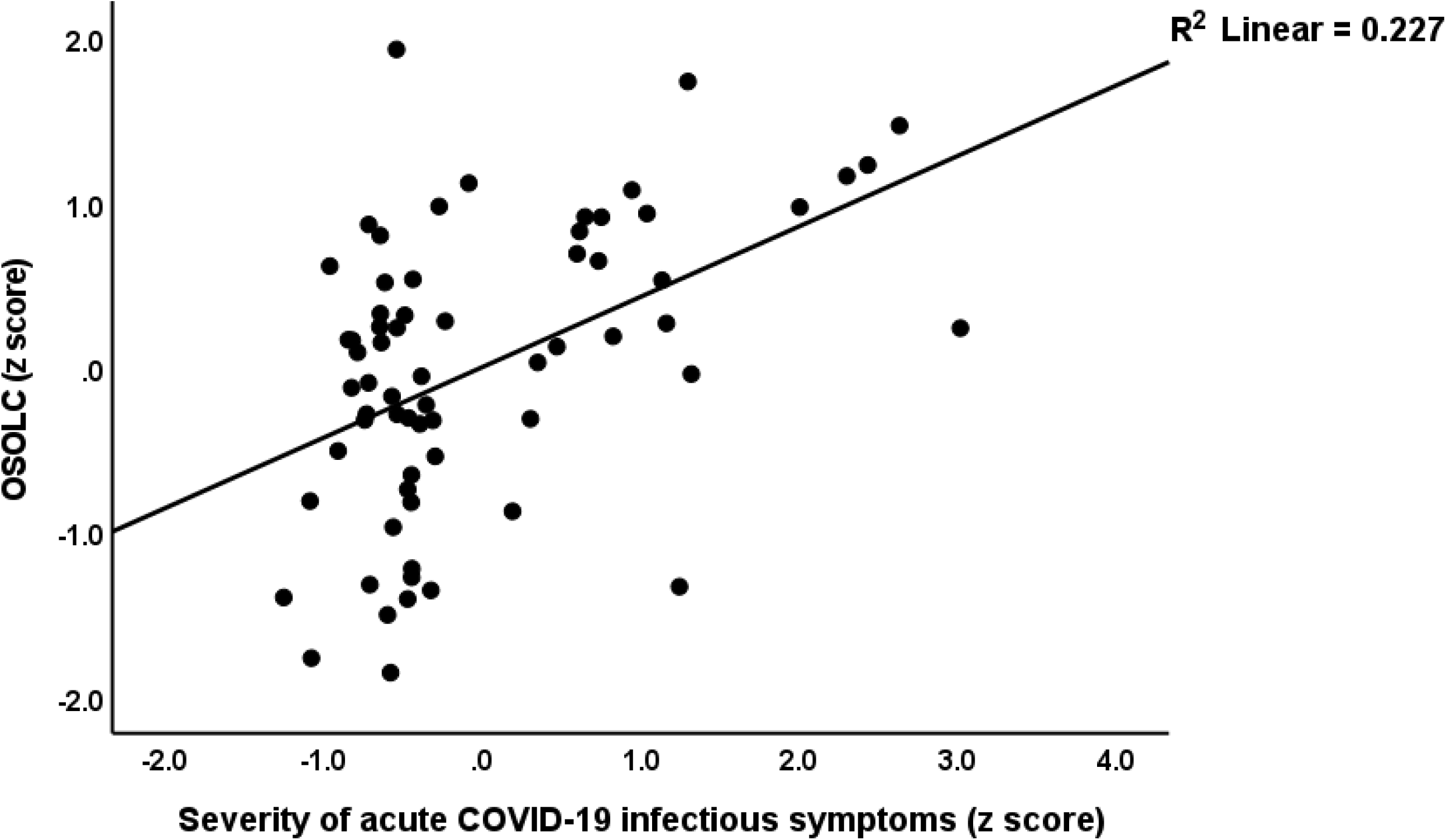
Partial regression of the overall severity of Long COVID (OSOLC) on the severity of COVID-19 infection symptoms (p<0.001).

**Figure 2.**
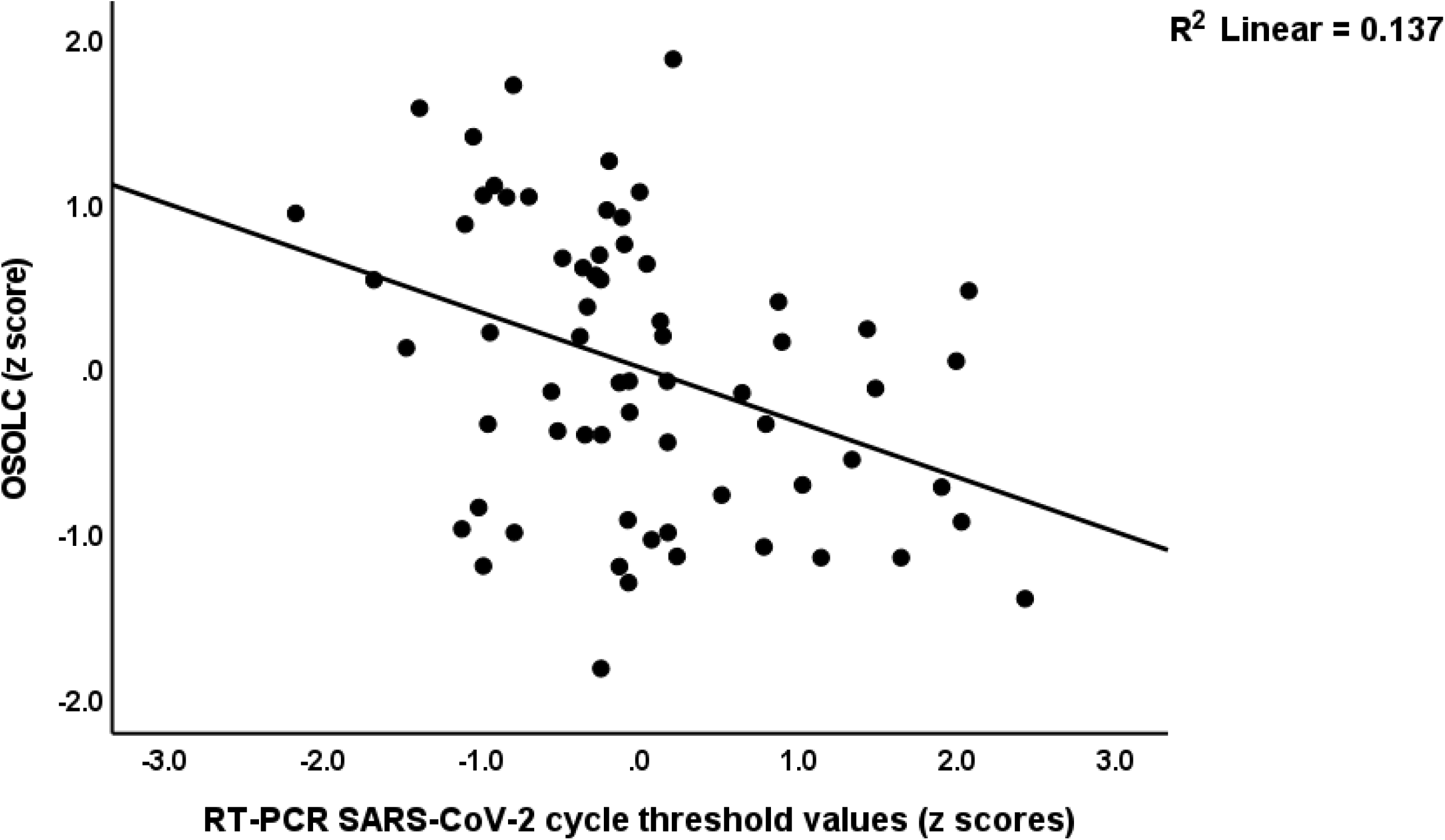
Partial regression of the overall severity of Long COVID (OSOLC) on the SARS-CoV-2 PCR cycle threshold values during the acute infectious phase (p<0.001).

## Discussion

### Clinical aspects of Long COVID

This study’s primary finding is that all five clinical subdomains of LC are significantly intercorrelated, and that we were able to extract a singular latent construct from the affective symptoms (depression, anxiety, vegetative symptoms, concentration difficulties), CFS/ME manifestations (chronic fatigue, fibromyalgia, headache, post-exertional malaise), respiratory symptoms (dry cough, dyspnea), and neurological symptoms (anosmia, ageusia, syncope, vertigo, tremors, paresthesia). Consequently, these symptom categories should be considered as manifestations of a shared underlying factor, specifically the physio-affective phenome of LC. The latter thus indicates the severity of LC symptoms.

The results align with other research indicating that a single latent construct can be derived from depressive symptoms, anxiety, CFS/ME symptoms, and physio-somatic symptoms (Al-Hakeim et al., 2023a; Almulla et al., 2024d; Maes et al., 2024). In this research, the physio-somatic symptoms were derived using three neuropsychiatric rating scales: the Hamilton Depression Rating Scale, the Hamilton Anxiety Rating Scale, and the Fibro-Fatigue scale. Therefore, the assessments of the physio-somatic symptoms were not entirely independent of depression and anxiety because the rating scales are constructed to show convergence validity. In the present study, we evaluated various symptoms from the WHO symptom list, ensuring that these symptoms were not chosen based on the convergence of rating scales.

Thus, the results of the current study show that the four symptom dimensions assessed here are manifestations of the physio-affective phenome of LC. The latter should be regarded as a continuum of severity of illness. Likewise, cluster analysis did not detect qualitatively distinct classes but merely classes that differ with respect to severity of the four dimensions. The findings of the above studies do not concur with previous views that LC may encompass various syndromes (Ely et al., 2024; Mahase, 2020; NIHR, 2020).

### The severity of the infectious phase predicts Long COVID

The second principal conclusion of this study is that the physio-affective phenome of LC and its subdomains and target symptoms such as post-exertional malaise are predicted by markers from the acute period of the illness. The most adequate predictor, as indicated by the standardized beta coefficients from the regression analyses, was the symptoms of the acute infectious period, specifically dyspnea and productive sputum. Similarly, prior research demonstrated that decreased SpO2 and elevated peak body temperature, both indicative of heightened inflammation during the acute phase of illness, are predictive of LC symptoms (Al-Hadrawi et al., 2022). The present study establishes a connection between the acute infectious phase of disease and LC symptoms by demonstrating that real-time PCR SARS-CoV-2 Ct levels measured during acute infection can predict LC symptoms several months later. The PCR method employed in the current study does not inherently indicate infectiousness, although these results may partially represent viral load (Bustin and Nolan, 2020; Han et al., 2021). Consequently, our data may indicate that a reduced number of cycles required to provide a positive PCR test, indicative of a greater viral load, could predict the overall severity of LC, including its depressive symptoms, CFS/ME symptoms, as well as respiratory and neurological manifestations. The positive correlation between neutrophil percentage and the depression score of LC further illustrates how immune activation is associated with these symptoms (Maes et al., 1992).

We also detected that a reduced anion gap during COVID-19 infection is positively associated with the severity of LC. Many patients with acute COVID-19 infection show alkalosis (61%) or acidosis (14%), and the latter patients have a worse outcome (Zemlin et al., 2023). A low anion gap may indicate lowered serum albumin because of an inflammatory response (Haber et al., 2023).

### Other biomarkers of Long COVID

Additional significant biomarkers indicative of LC severity includes diminished TRAP and NOx levels. It was previously reported that LC correlates with elevated serum levels of myeloperoxidase, advanced oxidation protein products (AOPP), lipid peroxidation biomarkers, and nitric oxide metabolites, suggesting heightened oxidative and nitrosative stress during LC (Al-Hakeim et al., 2023a; Al-Hakeim et al., 2023b). Moreover, the previous authors identified reduced antioxidant defenses (lowered serum glutathione peroxidase and zinc) in LC patients compared to controls (Al-Hakeim et al., 2023a). Nonetheless, in the current study, reduced TRAP was evaluated several months before the LC assessment and, consequently, predicted LC. Elevated serum levels of malondialdehyde provide significant predictive value in differentiating LC from asymptomatic post-COVID individuals (Stufano et al., 2023). Additional studies have similarly documented heightened oxidative stress in LC (Hofmann et al., 2023; Noonong et al., 2023). Consequently, it is plausible to conjecture that diminished TRAP, as a marker of total plasma antioxidant capacity, predisposed individuals to elevated oxidative stress during LC, resulting in damage to proteins and lipids. In individuals with moderate COVID-19, serum thiol groups were longitudinally correlated with LC over a 12-month period (Vlaming-van Eijk et al., 2024). Interestingly, we found that PON1 QR192 heterozygous individuals may show some protection against the more severe LC phenotype. The PON1 QR192 genotype and PON1 activities are associated with major depression (Moreira et al., 2019a; Moreira et al., 2019b).

A prior investigation identified elevated nitric oxide metabolites in a subset of patients with LC (Al-Hakeim et al., 2023a), while the present study found that reduced nitric oxide levels predicted LC some months later. Early SARS-CoV-2 is marked by diminished nitric oxide availability in association with disease severity (Montiel et al., 2022). It was previously proposed that a deficiency of nitric oxide could lead to adverse effects in COVID-19 patients (Nikolaidis et al., 2021). Significantly, low-dose inhaled nitric oxide may enhance blood oxygenation in critically ill patients suffering from acute lung injury and may provide advantages for COVID-19 patients with pneumonia (Brown, 2023; Ferrari and Protti, 2022). The reductions in nitric oxide bioavailability may originate from oxidative-related endothelial dysfunctions in severe COVID-19 patients (Ferrari and Protti, 2022). It is plausible that during the development of LC nitric oxide production increases due to diminished antioxidant defenses and heightened oxidative stress. The present investigation showed that reduced ALT levels predicted the depressive and neurological manifestations of LC. A meta-analysis of 23 studies indicated that some patients exhibit either decreased or elevated ALT readings (Bzeizi et al., 2021). It should be noted that, in COVID-19, ALT levels are inversely correlated with SARS-CoV-2 antigen (Rouka et al., 2021). The findings of an additional investigation emphasize that ALT levels are inversely correlated with increased death rates in COVID-19 (Goel et al., 2021). In the elderly and heart failure patients, diminished ALT levels correlate with heightened sarcopenia and frailty, characterized by fatigue, decreased gait velocity, and diminished survival (Segev et al., 2020; Vespasiani-Gentilucci et al., 2018).

## Limitations

While the study yielded sufficient power (minimal 0.8) to analyze the predictive power of the clinical and biomarkers of LC, the sample was rather small to perform cluster analysis, and, therefore, could have missed relevant subclasses. Furthermore, this study did not include all symptom domains such as skin disorders (rash, hair loss, itching), metabolic disease (obesity), insomnia (including sleep hypopnea or apnea), and hormonal changes (dysmenorrhea). Furthermore, it remains ambiguous if gastro-intestinal symptoms may be regarded as an additional manifestation of the first principal component reflecting the physio-affective phenome developed herein. The gastro-intestinal score exhibited a substantial correlation with the other four domain scores; nevertheless, this domain did not achieve a sufficiently high loading (specifically > 0.624, therefore below 0.7). The lower number of patients reporting gastrointestinal symptoms may have constrained the variability of its score, hence affecting its covariation with other symptom domains and the first principal component. In addition, the findings obtained in Thailand merit replication in other cultures and countries.

## Conclusions

The five LC subdomains examined here exhibit considerable intercorrelations. Therefore, LC may represent a clinically homogenous condition distinguished by closely interrelated affective, chronic fatigue syndrome, neurological, gastrointestinal, and respiratory subdomains. We were able to extract a singular latent concept that encompasses affective, CFS/ME, respiratory, and neurological symptoms, termed the physio-affective phenome of LC. The latter and its subdomains and target symptoms such as post-exertional malaise were significantly predicted by clinical symptoms and biomarkers indicative of the severity of the acute infectious COVID-19 stage, along with related inflammatory and oxidative stress biomarkers. LC is predicted by the severity of the acute infectious phase.

## Author’s contributions

All authors contributed to the work and approved the last version of the manuscript.

## Ethics approval and consent to participate

The research project (IRB number 114/64) was approved by the Institutional Review Board of Chulalongkorn University’s institutional ethics board. All patients gave written informed consent prior to participation in the study.

## Funding

This work was supported by the FF60 grant and Sompoch Endowment Fund from the Faculty of Medicine, MDCU to MM. CT, PS and NH were supported by the e-ASIA Joint Research Program (e-ASIA JRP) from the National Science and Technology Development Agency; Ratchadapisek Somphot Fund, Faculty of Medicine, Chulalongkorn University, grant number RA-MF 03/68.

## Conflict of interest

The authors have no commercial or other competing interests concerning the submitted paper.

## Data Availability Statement

The dataset that was made and/or analyzed during this study will be available from the corresponding author (MM) once it has been fully used by the authors and a reasonable request has been made.

## Notes

### Competing Interest Statement

The authors have declared no competing interest.

