## supplementary file for "The physio-affective symptoms of Long COVID are strongly predicted by the severity of the acute infectious phase, and lowered antioxidants, nitric oxide, and alanine transaminase levels"

**ELECTRONIC SUPPLEMENTARY FILE (ESF)**

ESF, Table 1. Long COVID symptoms assessed in the present study.

| Depressive symptoms | Depressed mood |
| --- | --- |
|  | Loss of interest |
|  | Anxiety |
|  | Loss of appetite |
|  | Weight loss |
|  | Forgetfullness |
|  | Concentration disorders |
|  | Impaired decision making |
|  | Insomnia |
|  | Slowness movements |
| CFS/ME symptoms | Fatigue |
|  | Post-exertional malaise |
|  | Headache |
|  | Muscle pain |
|  | Muscle stiffness |
| Respiratory symptoms | Pain on breathing |
|  | Dry cough |
|  | Shortness of breath |
|  | Chest pain |
| Neurological symptoms | Tremor |
|  | Numbness |
|  | Swallowing problems |
|  | Weakness in the limbs |
|  | Dizziness |
|  | Balance problems |
|  | Unsteady gait |
|  | Impairments of voluntary motor movements |
|  | Myoclonic jerks |
|  | Fainting |
|  | Seizures |
|  | Smell loss |
|  | Loss of taste |
|  | Hearing problem |
| Gastro-intestinal symptoms | Constipation |
|  | Diarrhea |
|  | Nausea |
|  | Stomach pain |
